# SARS-CoV-2 seroprevalence in healthcare workers of dedicated-COVID hospitals and non–COVID hospitals of District Srinagar, Kashmir

**DOI:** 10.1101/2020.10.23.20218164

**Authors:** Muhammad Salim Khan, Inaamul Haq, Mariya Amin Qurieshi, Sabhiya Majid, Arif Akbar Bhat, Muhammad Obaid, Tanzeela Bashir Qazi, Iqra Nisar Chowdri, Iram Sabah, Misbah Ferooz Kawoosa, Abdul Aziz Lone, Shahroz Nabi, Ishtiyaq Ahmad Sumji, Rafiya Kousar

## Abstract

**Background and objective:** SARS-CoV-2 infection poses tremendous challenge to the healthcare system of nations across the globe. Serological testing for SARS-CoV-2 infection in healthcare workers, which form a high-risk group, helps in identifying the burden of hidden infection in an institutional setting.

**Methods:** We present the results of a cross-sectional serosurvey in healthcare workers from two different hospital settings based on their role in the management of SARS-CoV-2 patients in District Srinagar, Kashmir. In addition to testing for the presence of SARS-CoV-2 specific IgG, we collected information on influenza-like symptoms in the last four weeks and the status of RT-PCR testing. SARS-CoV-2 specific IgG antibodies were detected in serum samples using a sensitive and specific chemiluminescent microparticle immunoassay technology.

**Interpretation and Conclusion:** Of 2915 healthcare workers who participated in the study, we analysed data from 2905 healthcare workers. The overall prevalence of SARS-CoV-2 specific IgG antibodies was 2.5% (95% CI 2.0-3.1) in the healthcare workers of District Srinagar. Healthcare workers who had ever worked at a dedicated-COVID hospital had a substantially lower seroprevalence of 0.6% (95% CI: 0.2 - 1.9). Among healthcare workers who had tested positive for RT-PCR, seroprevalence was 27.6% (95% CI: 14.0 - 47.2).The seroprevalence of SARS-CoV-2 infection in healthcare workers of District Srinagar is low, reflecting that a high proportion of healthcare workers are still susceptible to the infection. It is crucial to lay thrust on infection prevention and control activities and standard hygiene practices by the healthcare staff to protect them from acquiring infection within the healthcare setting.

## Introduction

India is emerging as the world’s biggest hotspot for severe acute respiratory syndrome coronavirus 2 (SARS-CoV-2) infection, second only to the United States of America, with more than four million recorded infections in seven months following the first reported case on 30^th^ January 2020[1, 2]. Kashmir, a northern territory of India, reported its first positive case of novel coronavirus on 18^th^March 2020 from a central district Srinagar which is currently the most affected place within the territory from where presently more than 20% of new cases are reported each day[3]. In addition to three tertiary care hospitals, one District Hospital, and twoSub-district Hospitals in District Srinagar, Primary Health Centres sizeably caters to the healthcare needs of the people. They are playing a pivotal role in the current pandemic for containment of SARS-CoV-2 infection. The healthcare workers, because of their job profile, are at a higher risk of acquiring SARS-CoV-2 infection[4–6]. As the proportion of individuals with asymptomatic infection is relatively high and variable across different regions, there is growing evidence which suggests that this hidden pool is serving as a potential source of infection for the general population and health care workers do not behave differently[7, 8].

Many countries have started testing for the presence of antibodies against SARS-CoV-2 infection, both at the population level and in specific groups like healthcare workers. Seroepidemiological studies are crucial in understanding the dynamics of SARS-CoV-2 infection. Seroepidemiological studies have been conducted in many places among the general population, but there is insufficient data on healthcare workers. World Health Organisation, in its scientific briefing on 24^th^ April, encouraged the member states to conduct seroepidemiological studies in the context of coronavirus disease (COVID-19) for a better understanding of the extent of infection[9–11].

Therefore, to find out the prevalence of SARS-CoV-2 infection among healthcare workers, we undertook this seroepidemiological study by testing for SARS-CoV-2 specific IgG to get an insight into the extent of infection among healthcare workers in our setting.

## Material and Methods

### Study design and settings

We conducted this seroepidemiological study to find out the presence of IgG antibodies against SARS-CoV-2 among healthcare workers of District Srinagar, Kashmir. We commenced the data collection from 15^th^June 2020 and completed it in two weeks. As a part of the preparedness of the health system for the pandemic, the central Ministry of Health and Family Welfare of the country specified guidelines for health facilities for the management of COVID-19 cases. Thus, hospitals were categorized into dedicated-COVID hospitals and non-COVID hospitals.

Dedicated-COVID hospitals provide comprehensive care to COVID-19 patientsexclusively, with round the clock facility of fully functional intensive care units, ventilators, and beds with assured oxygen support [12].

### Participants

We invited healthcare workers from threededicated-COVID, seven non-COVID tertiary care hospitals, two sub-district hospitals, and four primary health centresacross the District (Figure 1). Administrative heads of the hospitals received written communication for permission to conduct the study and invite all healthcare workers in their hospitals for participation. Participation in the study was also encouraged through use of social media and word of mouth. All frontline healthcare workers,including doctors, administrative and laboratory personnel, technicians, field workers who were involved in surveillance activity, and other supporting staff, were part of the study. The participation of healthcare workers in the survey was voluntary.

**Fig I:**
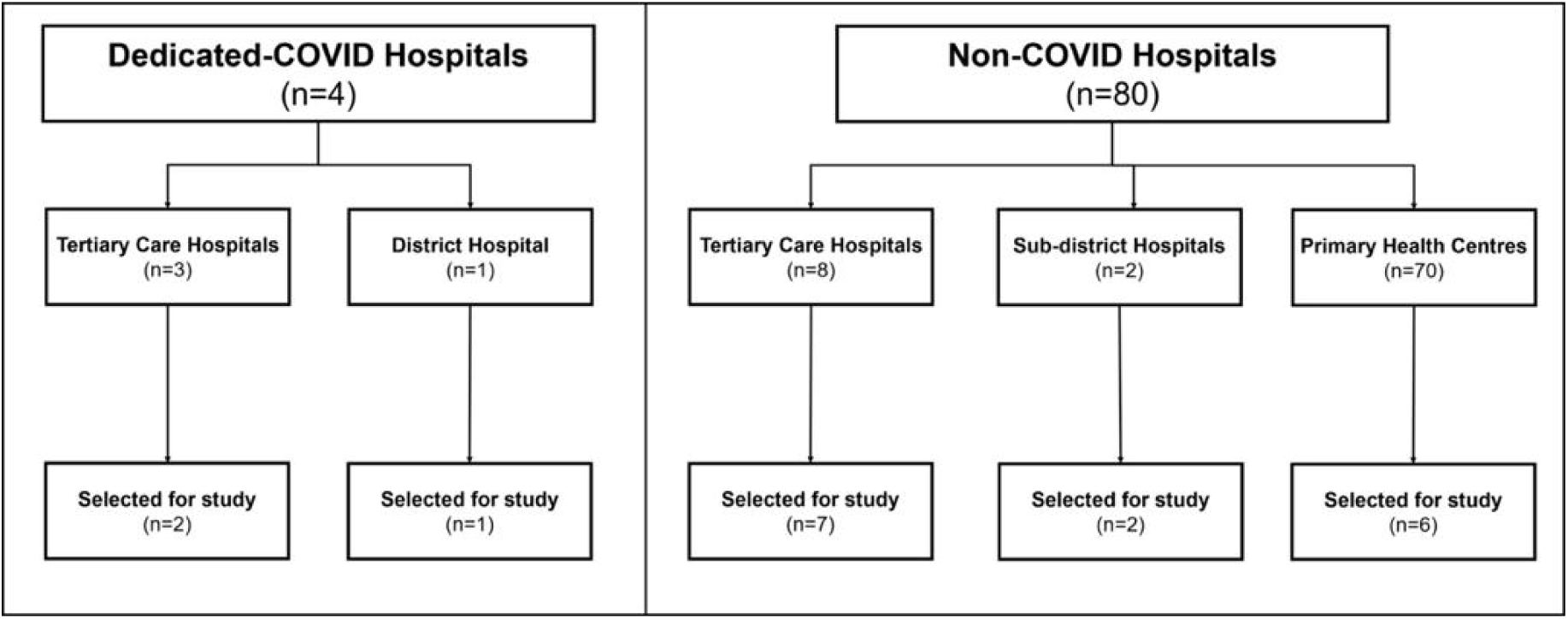
Health facilities in District Srinagar and the number of hospitals selected for the study.

### Procedure

We gathered all the information on a form generated on mobile phones using Epicollect5, a free mobile phone data gathering tool widely used in public health research for the collection of large scale data[13].Doctors, specifically trained in the use of Epicollect5, conducted the interviews.We collected information about the participants’ role in the current pandemic in terms of their involvement in providing care to COVID-19 patients. Participants also provided information on risk factors for SARS-CoV-2 infection (history of travel since 1^st^ January 2020, flu-like symptoms in the last four weeks). Following the interview, trained phlebotomists collected 3-5mL of venous blood under aseptic precautions. Centrifugation of the samples was done at the facility. The centrifuged samples were transported to a central laboratory for further processing and testing. Centrifugation was done at the central laboratory for sites that did not have the facility. Standard operating procedures were strictly adhered to during collection, transportation, and testing of blood samples.

### Variables

Our primary outcome of interest was the presence of IgG antibodies against SARS-CoV-2 among healthcare workers.

### Laboratory procedure

We used a chemiluminescent microparticle immunoassay (CMIA) based procedure for the detection of SARS-CoV-2 specific IgG antibodies in serum samples. We followed the manufacturer’s recommendations for the process. The assay is an automated, two-step immunoassay for the qualitative detection of IgG antibodies to SARS-CoV-2 in human serum and plasma. The test result was considered positive for SARS-CoV-2 IgG if the ‘index value’ was ≥1.4 as provided by the manufacturer[14].

Laboratory data was entered in a similar fashion using Epicollect5. Two trained persons independently entered the laboratory results in two separate forms.Data from the two forms were checked for any discrepancies, in which case, the source data was referred to for necessary corrections. The information gathered during the interview and the laboratory results were linked with the help of a unique identification number, which was generated at the time of the interview.

### Statistical analysis

We estimated the proportion and 95% confidence interval for the SARS-CoV-2 specific IgG antibody in healthcare workers. We looked for the difference in seroprevalence across gender, age group, specific occupation group, and type of health facility (dedicated-COVID hospital versus non-COVID hospital). Seroprevalence was also estimated separately for healthcare workers who reported symptoms in the last four weeks, had a history of exposure to a known case of SARS-CoV-2 positive patient, or had undergone testing by reverse transcriptase-polymerase chain reaction (RT-PCR). We used the Chi-square test to report two-sided p-values for comparison of seroprevalence between groups. The exact test was used instead of the Chi-square test when the expected frequency was less than five in more than 20% of the cells. We did not adjust p-values for multiple comparison. Stata version 15.1 was used for data analysis.

### Ethics approval and consent to participate

The Institutional Ethics Committee of Government Medical College Srinagar approved the study (Ref. No. 1003/ETH/GMC dated 13-05-2020). We obtained written, informed consent from all participants.

## Results

Out of 7346 healthcare workers in the District 2915 participated in the study. The overall response rate was 40%, 47% among doctors, and 37% among paramedical and other hospital staff. We analysed the information gathered from 2905 healthcare workers. The information on the Epicollect5 interview form was missing for nine individuals, and the laboratory report was missing for one. The mean age of the participants was 38.6 years, and 35.8% were females. One third (33.8%) of the participants were doctors, of whom nearly half were resident doctors. (Table 1)

**Table I:**
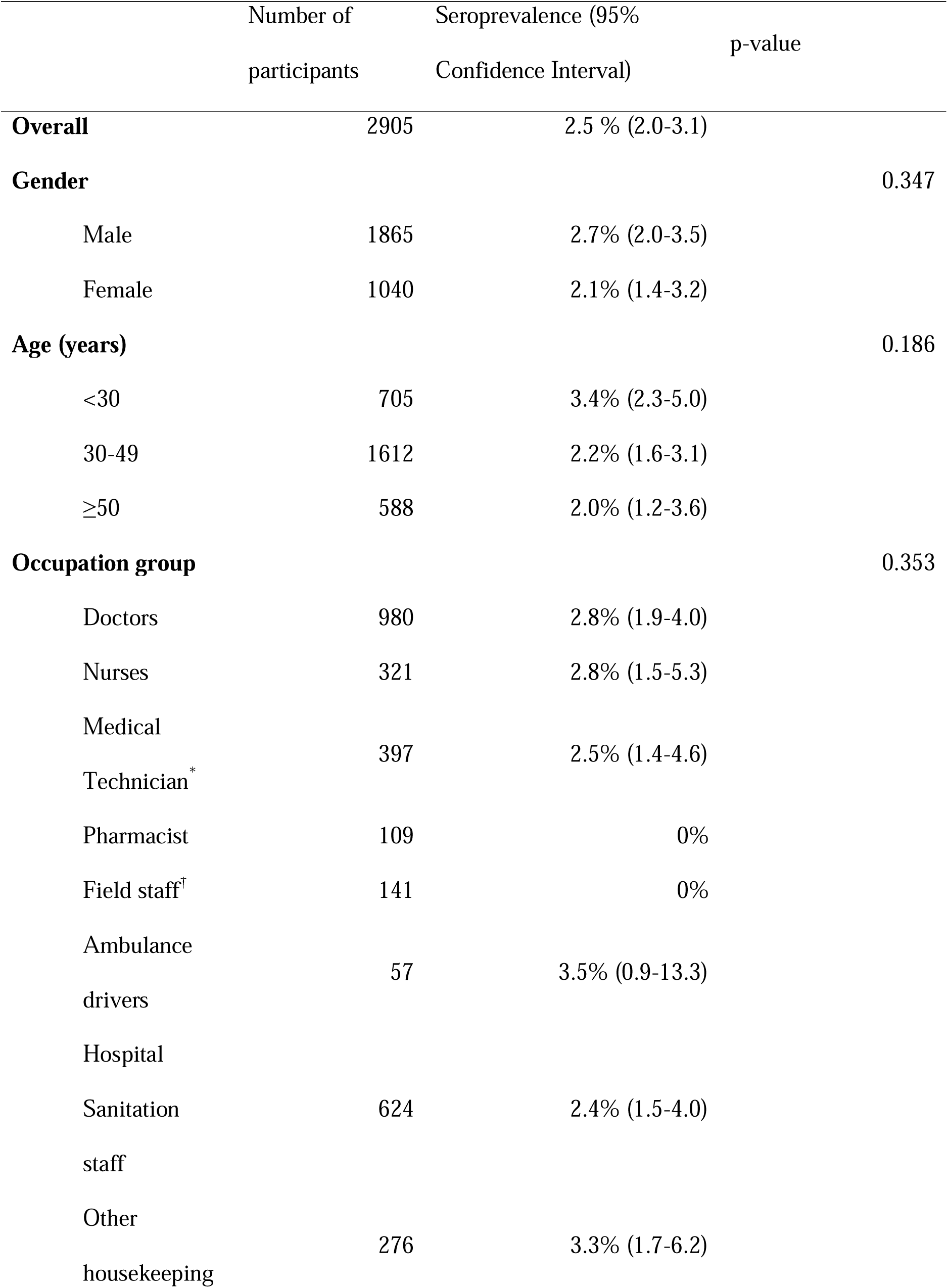
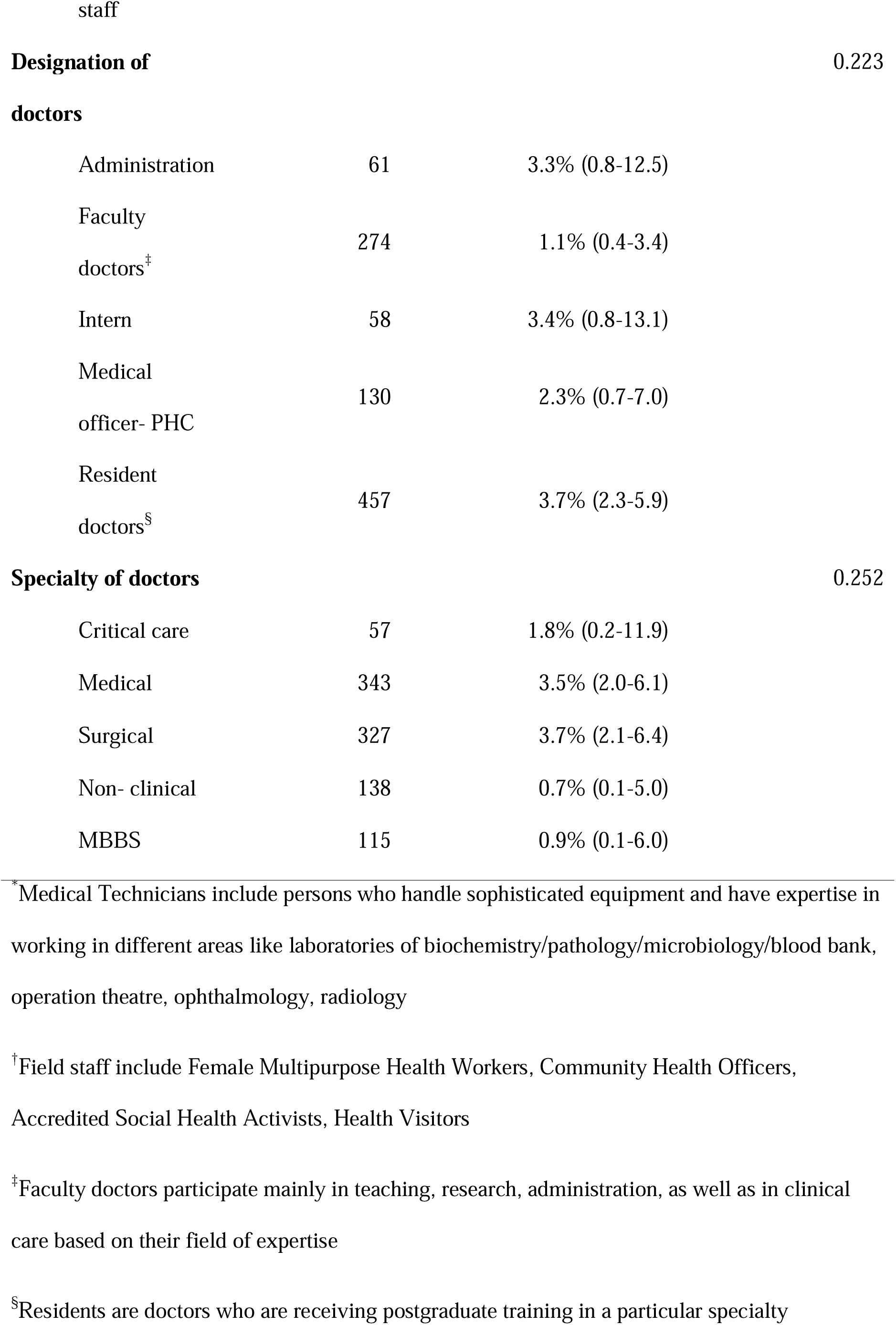
Seroprevalence of SARS-CoV-2 specific IgG antibodies across baseline characteristics of healthcare workers

Of the 2905 healthcare workers, 123 (4.2%) reported an influenza-like illness(fever and cough) in the four weeks preceding the interview, and 339 (11.7%) reported having close contact with a coronavirus disease (COVID-19) case. Seven hundred sixty (26.2%) healthcare workers had taken an RT-PCR, and out of these, 29 (3.8%) had a positive test result. (Table 2)

**Table II:** Seroprevalence of SARS-CoV-2 specific IgG antibodies by clinical characteristics and specific risk factors

|  | Number of participants | Seropositivity (95% CI) | p-value |
| --- | --- | --- | --- |
| **Symptoms of an influenza-like illness\*** |  |  | <0.001 |
| Present | 123 | 12.2% (7.4-19.3) |  |
| Absent | 2782 | 2% (1.6-2.6) |  |
| **RT-PCR status** |  |  | 0.390 |
| Ever done | 760 | 2.9% (1.9-4.4) |  |
| Never done | 2145 | 2.3% (1.8-3.1) |  |
| **RT-PCR result(n=760)** |  |  | <0.001 |
| Positive | 29 | 27.6% (14.0-47.2) |  |
| Negative | 731 | 1.9% (1.1-3.2) |  |
| **Ever put under quarantine** |  |  | 0.009 |
| Yes | 268 | 4.9%(2.8-8.2) |  |
| No | 2637 | 2.2%(1.7-2.9) |  |
| **Ever worked at a quarantine centre** |  |  | 0.667 |
| Yes | 60 | 3.3% (0.8-12.7) |  |
| No | 2845 | 2.5% (2.0-3.1) |  |
| **Ever worked in a dedicated-COVID hospital** |  |  | 0.004 |
| Yes | 480 | 0.6% (0.2-1.9) |  |
| No | 2425 | 2.8% (2.3-3.6) |  |
| **History of close contact** |  |  | 0.014 |
| **with a COVID-19 case** |  |  |  |
| Yes | 339 | 4.4% (2.7-7.2) |  |
| No | 2566 | 2.2% (1.7-2.9) |  |
| **History of travel after 1<sup>st</sup>** |  |  | 0.963 |
| **January 2020** |  |  |  |
| Yes | 165 | 2.4% (0.9-6.3) |  |
| No | 2740 | 2.5% (2.0-3.1) |  |

\* A history of fever and cough with or without other symptoms, within four weeks preceding the date of interview

We reported an overall seroprevalence of 2.5% (95% CI 2.0-3.1) for SARS-CoV-2 specific IgG antibodies among the healthcare workers of District Srinagar. Seropositivity was not significantly related to age, gender, or occupation group of healthcare workers. (Table 1)

We found a significantly higher seropositivity rate among those with a history of influenza-like illness (p<0.001),a history of positive RT-PCR (p<0.001), ever put under quarantine (p=0.009), and those with a self-reported history of close contact with a COVID-19 case (p=0.014). Surprisingly, healthcare workers who had ever worked at a dedicated-COVID hospital had a low prevalence of infection in comparison to those who never worked at a facility dedicated to such patients (p=0.004).

## Discussion

We aimed to estimatethe seroprevalence of SARS-CoV-2 infection in healthcare workers working in different hospital settings of District Srinagar, Kashmir. In general, seroprevalence was low (2.5%), with little difference across gender or occupation group.

Seroprevalence studies done in healthcare workers in different settings have revealed varied findings with estimates ranging from 1% to 10.2%[15–18]. Generally speaking, the seroprevalence among healthcare workers is not disproportionate when compared to the general population, which reflects the different dynamics of this infection when compared to other infections in healthcare settings. The low seroprevalence observed in our study coincides with the overall low infection rate in the population. During the study period, the District reported a median daily number of new infections of 28 (IQR 17-46),which is indicative of the early phase of the pandemic in the population at the time of study[19].

Among the healthcare worker occupation groups, ambulance drivers, and housekeeping staff of the hospitals reported the highest seroprevalence. The possible explanation could be a lower level of standard hygiene practices and inadequate use or reuse of protective gear due to a shortage of personal protective equipment and lack of training on donning and doffing of personal protective equipment[20].

Interestingly, healthcare workers who were working in a dedicated-COVID hospitals or had ever worked there had a very low seroprevalence, 0.6% (95%CI 0.2-1.9). As these facilities were dedicated to the management and care of SARS-CoV-2 patients, the hospital staff was taking extreme precautions while providing care to these patients. These hospitals were on high alert and were strictly adhering to infection prevention and control practices. Secondly, being a potentially high-risk area for transmission of infection, these facilities did staff rationing to reduce the duration of exposure in the healthcare staff. Staff from other hospitals, which included resident doctors, nurses, and other support staff, were put on rotational duty at the dedicated-COVID hospitals to compensate for the additional requirement. We did not, however, ascertain the adherence to infection prevention and control guidelines in the two different categories of hospitals. Based on the level of exposure in healthcare workers of University Hospital in Germany, Kortha J et al. reported a lower seroprevalence in the high-risk group, which included hospital staff who come in daily contact with SARS-CoV-2 patients and work in intensive care units[16].

We report a two-fold higher seroprevalence (4.4%) in healthcare workers who reported close contact with a SARS-CoV-2 positive patient as compared to those who did not report any such contact. Respiratory infections pose a greater health risk in an occupational setting to healthcare workers. There is no classic literature available that has established the risk factors for transmission for the SARS-CoV-2 infection. So far, globally,among thousands of healthcare workers infected with SARS-CoV-2 infection, close contact with a SARS-CoV-2 patient has been identified as one of the leading risk factors, among others which include lack of personal protective equipment, poor infection prevention and control practices, work overload, and pre-existing health condition[21].

The seroprevalence estimates in healthcare workers who reported symptoms suggestive of SARS-CoV-2 infection in the preceding four weeks was 12.2%,suggesting the presence of other circulating respiratory pathogens that give rise to these symptoms.

Among those who reported a previously positive RT-PCR for SARS-CoV-2, only 27.6% showed the presence of SARS-CoV-2 specific IgG antibodies. The inability to mount an antibody-mediated immune response or early conversion to seronegative status during the convalescence phase has been suggested[22]. On the contrary, seropositivity in previously negative RT-PCR subjects was 1.9 %. There are few plausible explanations for such observations. Firstly, several studies have reported 2%-29% false-negative results from RT-PCR[23–25]. Secondly, the sensitivity of the SARS-CoV-2 assay is influenced by the timing of test post symptoms or RT-

PCR positivity. The sensitivity of the test assay we used varies from 53.1% at day-7 to 100% at day-17 post-infection[26].Variable viral load and difference in duration of viral shedding are the other possible reasons for false-negative RT-PCR[27, 28].

In our study, seroconversion among asymptomatic healthcare workers who tested positive on RT-PCR was 20.8%. Among healthcare workers who reported influenza-like illness and were RT-PCR positive, seropositivity was 60%. In a study from a Medical University Hospital in China, 40% and 13% asymptomatic and symptomatic SARS-CoV-2 positive cases, respectively, became seronegative, after initial seropositivity, eight weeks after hospital discharge[29].

Serological testing gives an insight into the immune status of the healthcare workers against SARS-CoV-2 infection. However, it is premature to conclude that those who have developed antibodies against the infection are risk-free or protected against reinfection. As the epidemic progresses, more and more healthcare workers are likely to get infected. Serial cross-sectional serosurveys can helpin monitoring the progression of the pandemic within a healthcare setting and guide the hospital authorities in resource allocation.

### Strengths

We used Abbott Architect SARS-CoV-2 IgG assay, which has exhibited a high level of consistency and performance characteristics when tested on the different patient populations. The participation rate was reasonably good in our study, and we included all the major hospitals in the District. The findings from our study could be considered as being representative of the healthcare workers in the District.

### Limitations

We did not collect information on the timing of symptoms and the date at which the healthcare workers became RT-PCR positive.With a cross-sectional study design, we cannot ascertain the reconversion from initial IgG positive to IgG negative status, which warrants a cohort study.

Participation in the survey was voluntary. There is thus a possibility of selection bias. Some healthcare workers with recent exposure or those who were symptomatic at the time of the study might not have participated, thinking that they would not benefit by IgG testing in the early phase of infection.

## Conclusions

We conclude that the seroprevalence of SARS-CoV-2 infection was low among the healthcare workers of District Srinagar at the time of the study. Healthcare workers in a dedicated-COVID hospital or who had ever worked in such a facility had lower seroprevalence suggesting adherence to standard operating procedures while dealing with SARS-CoV-2 patients. It is crucial to lay thrust on infection prevention and control activities in the hospital settings.

Training and re-training of sanitation and other housekeeping staff about standard hygienic practices and appropriate use of the protective gear are needed.

## Data Availability

THE DATA WILL BE MADE AVAILABLE ON REQUEST

## List of abbreviations

SARS-CoV-2: Severe acute respiratory syndrome coronavirus 2
COVID-19: Coronavirus disease
CMIA: Chemiluminescent microparticle immunoassay
CI: Confidence interval

## Data Availability statement

Data shall be made available on request through corresponding author.

## Conflict of interest

Authors declare that they have no competing interests

## Funding statement

The study received mainly institutional funding from Government Medical College Srinagar with support from District Disaster Management Authority Srinagar. The funding bodies have no role in design, collection, analysis, interpretation, or writing of manuscript.

## Acknowledgments

We are thankful to Principal and Dean Government Medical College, Srinagar, Professor Samia Rashid, and District Commissioner, Srinagar, Shahid Iqbal Chowdhary for their support. We acknowledge the support rendered by the Directorate of Health Services, Kashmir, Chief Medical Officer, Srinagar, Block Medical Officers, and Zonal Medical Officers of District Srinagar, Kashmir. We highly appreciate the efforts put in by medical interns in data collection and laboratory in-charge Gulzar Ahmad Wani, Ph.D. scholar, Biochemistry, and his staff, who were involved in this study. We thank the study participants for understanding the importance of this study and giving their time and consent for participation.

## Notes

### Competing Interest Statement

The authors have declared no competing interest.

### Author Declarations

IT IS AN OBSERVATIONAL STUDY INTSTITUTIONAL ETHICAL COMMITTEE GMC SRINAGAR

